# Dosimetry of [^64^Cu]FBP8: a fibrin-binding PET probe

**DOI:** 10.1101/2024.06.27.24309589

**Authors:** David Izquierdo-Garcia, Pauline Désogère, Anne L. Philip, David E. Sosnovik, Ciprian Catana, Peter Caravan

## Abstract

**Purpose:** This study presents the biodistribution, clearance and dosimetry estimates of [^64^Cu]Fibrin Binding Probe #8 ([^64^Cu]FBP8) in healthy subjects.

**Procedures:** This prospective study included 8 healthy subjects to evaluate biodistribution, safety and dosimetry estimates of [^64^Cu]FBP8, a fibrin-binding positron emission tomography (PET) probe. All subjects underwent up to 3 sessions of PET/Magnetic Resonance Imaging (PET/MRI) 0-2 hours, 4h and 24h post injection. Dosimetry estimates were obtained using OLINDA 2.2 software.

**Results:** Subjects were injected with ∼400 MBq of [^64^Cu]FBP8. Subjects did not experience adverse effects due to the injection of the probe. [^64^Cu]FBP8 PET images demonstrated fast blood clearance (half-life = 67 min) and renal excretion of the probe, showing low background signal across the body. The organs with the higher doses were: the urinary bladder (0.075 vs. 0.091 mGy/MBq for males and females, respectively); the kidneys (0.050 vs. 0.056 mGy/MBq respectively); and the liver (0.027 vs. 0.035 mGy/MBq respectively). The combined mean effective dose for males and females was 0.016 ± 0.0029 mSv/MBq, lower than the widely used [^18^F]fluorodeoxyglucose ([^18^F]FDG, 0.020mSv/MBq).

**Conclusions:** This study demonstrates the following properties of the [^64^Cu]FBP8 probe: low dosimetry estimates; fast blood clearance and renal excretion; low background signal; and whole-body acquisition within 20 minutes in a single session. These properties provide the basis for [^64^Cu]FBP8 to be an excellent candidate for whole-body non-invasive imaging of fibrin, an important driver/feature in many cardiovascular, oncological and neurological conditions

## Introduction

Fibrin is an insoluble blood protein formed from fibrinogen as a result of the coagulation cascade and is one of the main components of thrombus (blood clot). Fibrin occurs in thrombosis, wound healing and in certain pathologies but is absent in blood plasma or healthy tissue. The presence of fibrin in pathology and its absence in healthy tissue implies that a fibrin-targeted imaging probe should have both high sensitivity and specificity for detecting disease if the background signal is low.

The identification of thrombus presence and location is crucial to improve patient outcomes. Current clinical practice utilizes different imaging modalities depending on vascular territory: contrast enhanced computed tomography (CT) for pulmonary embolus detection, transesophageal echo cardiography (TEE) for cardiac thrombi, duplex ultrasound for deep vein thrombosis. However each has its limitations: contrast enhanced CT is contraindicated in patients with poor renal function [1] and pregnant women [2], TEE requires sedation [3], duplex ultrasound cannot be performed in patients in casts [4], and none of these techniques can be used to examine multiple vascular territories. Molecular imaging of fibrin with positron emission tomography (PET) offers the potential to overcome these limitations and also provide molecular characterization of the thrombus to distinguish acute vs. chronic thrombus [5].

Extravascular fibrin is increasingly recognized to be implicated in other pathologies. In multiple sclerosis, extravascular fibrin has been proposed as a driver of neuroinflammation [6]. In Alzheimer’s disease, extravascular fibrin is linked to higher inflammation, neuronal loss and cognitive decline [7]. Extravascular fibrin is also present in most solid tumors [8]. In the lungs, the presence of extravascular fibrin is associated with acute respiratory distress syndrome [9], pulmonary fibrosis [10], and long-covid or chronic fatigue syndrome [11]. The use of a highly specific fibrin-binding probe together with the large field of view capability of PET provides a unique platform to quantify fibrin throughout the brain and body.

Utilizing a short cyclic peptide that has high affinity for fibrin and specificity for fibrin over fibrinogen [12], we synthesized and evaluated several fibrin-specific PET probes [13–16], and ultimately identified [^64^Cu]FBP8 as a candidate for clinical development [17–19]. We recently reported on the sensitivity and specificity of [^64^Cu]FBP8-PET for the detection of left atrial appendage thrombus in patients with atrial fibrillation [20]. [^64^Cu]FBP8-PET has also been shown to detect lung injury in patients with idiopathic pulmonary fibrosis [21]. There are also three active clinical trials using [^64^Cu]FBP8-PET on Clinicaltrials.gov, NCT05336695, NCT04022915, and NCT03830320. In light of the increasing use of [^64^Cu]FBP8, a more in-depth characterization of the probe is required. In this study, we present more details about the biodistribution, clearance and dosimetry estimates of [^64^Cu]FBP8 in healthy subjects. Radiation dosimetry studies are required for newly developed probes to provide estimates for the potential use of the probe in the clinic. This is of particular interest for [^64^Cu]FBP8 considering the likelihood of its potential repeated use in the clinic in high-risk cardiovascular subjects [22] and other important clinical areas as described above.

## Materials and Methods

### Synthesis of [^64^Cu]FBP8

[^64^Cu]FBP8 was synthesized with a chemical purity typically >99% as previously described [17, 19]. The probe consists of a 6-amino-acid cyclic peptide with a high affinity for fibrin, conjugated to a [^64^Cu]-containing chelator (NODAGA) as previously shown [20].

### Patient recruitment and safety monitoring

This study was approved by the Mass General Brigham (formerly Partners) Institutional Review Board (protocol number 2015P002385) and registered at clinicaltrials.gov (NCT03830320). Informed consent was obtained from all individual participants included in the study. A total of eight healthy volunteers (2 females, 6 males) aged 35±16 years, average BMI 24.7±4.8 Kg/m^2^ were recruited for this study. Simultaneous PET and MRI (3T) images were acquired on a Biograph mMR scanner (Siemens Healthcare, Erlangen, Germany). One subject (subject 3) could not complete the study due to a scanner failure during the acquisition and therefore was removed from the data analysis. Patient demographics are summarized in Table 1 below.

**Table 1:**
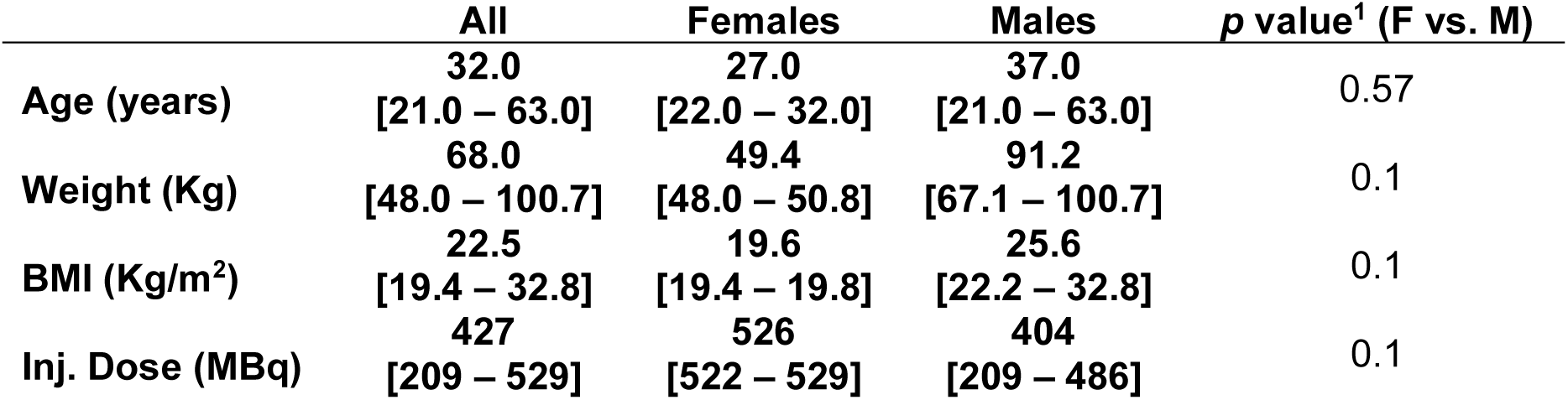
Subject Demographics and Injected Dose. Data presented as Median [min – max]. ^1^Wilcoxon Rank Sum Test.

Safety was monitored by direct communication with the subject and by recording vital signs before and after scan (respiratory rate, pulse, blood pressure, and temperature) and autonomic measures (heart rate, respiration rate, heart rhythm and pulse oximetry) before and after [^64^Cu]FBP8 injection, and throughout the scan. Electrocardiograms (ECG) were recorded before injection and at the end of the scans. Blood was drawn prior to [^64^Cu]FBP8 injection and post-imaging, and coagulation parameters were compared (Fibrinogen, PTT, PT/INR). Complete blood count (CBC) and a clinical chemistry panel were drawn and compared prior to [^64^Cu]FBP8 injection and 24 hours later for those healthy subjects who elected to come in for optional scans. A study physician followed subjects up with phone calls on the days after imaging sessions.

### PET/MR image acquisition

Subjects were asked to be imaged on a simultaneous PET/MRI scanner in three sessions over two days. On day 1, subjects were scanned continuously for the first 2 hours following injection of ∼350 MBq of [^64^Cu]FBP8 [18–19], followed by a break of 2 hours before undergoing a further scan. Subjects were asked to return the following day for another imaging session at ∼24 h post injection. PET emission data was acquired using a multi-bed whole-body acquisition protocol covering from top of the head to mid-thigh (in 5 bed positions). The first time-point was centered on the heart and lungs area, then moving down until mid-thigh, then to the top of the head and down to the neck. During the break time between sessions, subjects were asked to void their bladder. A total of seven whole-body time-points (frames) were acquired: 0, 25, 50, 75, 100, 240 and 1440 (24 h) min post injection. PET data at each bed position was acquired for 4 min. Four subjects (2 female, 2 male) completed all three sessions while the remaining 3 subjects (males) refused to complete the study after the first imaging session.

PET images were reconstructed in 3D mode, using Ordinary Poisson Ordered-Subset Expectation-Maximization (OP-OSEM) with 3 iterations and 21 subsets with a post-reconstruction Gaussian filter of 4mm isotropic, with corrections applied for normalization, photon attenuation, scatter, dead-time, randoms and sensitivity. Photon attenuation was corrected using the standard Dixon-based sequence approach provided by the manufacturer [23]. Images were reconstructed into a 256x256x127 matrix with voxel size 2.8x2.8x2.03mm^3^, for a total extended axial coverage of ∼1m.

Simultaneously, T2-weighted STIR HASTE images were acquired coronally during 2 concatenated breath-holds, using the following parameters per bed position: TE/TR = 0.1/1 s; TI = 0.2 s; flip angle = 120 degrees; reconstruction matrix = 256x179x36 voxels, 1.6x1.6x7mm^3^; total time of acquisition = 36s. Additionally, dual-echo Dixon images were acquired with the following parameters: TE = 1.2/2.5 ms; TR = 3.6 ms; flip angle = 10 degrees; reconstruction matrix = 192x126x128 voxels, 2.6x2.6x2.2 mm^3^; total acquisition time = 19 s.

### Biodistribution and Pharmacokinetics

PET data was converted into percentage of injected dose per milliliter (%ID/mL) by dividing by the injected activity. Time activity curves (TACs) were plotted to evaluate probe distribution in different organs. Additionally, PET data was also converted into standardized uptake values (SUV) to compare different organ uptake at 90 and 240 min post injection.

Blood pharmacokinetics was calculated from the PET images using a region of interest (ROI) measured on the center of the left ventricle by fitting to a biexponential function:

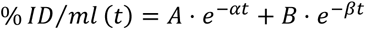

where the distribution half-life is defined as Ln(2)/α and elimination half-life as Ln(2)/β.

### Dosimetry Analysis

Human dosimetry estimates were calculated using the Organ Level Internal Dose Assessment (OLINDA) software v2.2 (OLINDA/EXM, Vanderbilt University, 2012) using the International Commission on Radiation Protection 89 (ICRP-89) male and female phantoms and using the ICRP-103 standard for organ weights [24]. We assumed uniform distribution of probe concentration throughout the organs. Sixteen ROIs were manually drawn [25–26] on the T2-weighted (HASTE) images using OSIRIX v4.1 (Pixmeo SARL, Geneva, Switzerland) covering all the organs with visible uptake above the background plus the most radiation-sensitive organs: brain, lungs, blood pool (left ventricle), heart (myocardium), breast (only females), pancreas, stomach, small intestine, large intestine (left colon), spleen, liver, gall bladder, kidneys, lumbar vertebrae (red marrow), bladder and soft-tissue (positioned on upper leg muscle). Additionally, for males ROIs in prostate and testes and for females ROIs in uterus and ovaries were added for a total of 18 ROIs per individual subject. ROIs were drawn on the coronal planes to cover most of the anatomical shape of the organ and, in all cases at least 3 consecutive planes (for larger organs) were provided [25]. ROIs were individually drawn on each acquired whole-body frame to avoid patient motion bias in the ROI analysis. ROIs were then propagated into the corresponding PET frames. Percent-injected dose per organ (%ID/organ) values were obtained by multiplying the %ID/mL values in each ROI by the phantom organ mass [24].

To account for differences in subject-specific body weight with respect to the male/female phantom potentially inducing a bias, %ID/organ values were scaled by the ratio of the actual subject’s weight over the male/female phantom weight from OLINDA to produce organ-normalized activities [26–27]. The time-dependent curves of %ID/organ were then fit using an in-house script (Python v.3.1) to provide the corresponding estimates of the organ time-integrated activity coefficients (TIAC), by analytical integration of a function using a combination of mono- and bi-exponentials [26].

TIACs for the urinary bladder were calculated using the voiding model on the OLINDA software in a similar way as explained in [28]. The model defines an excretion fraction and the half-life of the total urine content (see Supplementary material). We chose a typical 1h voiding interval which would be closer to the clinical situation, where patients would be asked to void prior to imaging after ∼1h post injection interval for radiotracer distribution.

Lumbar vertebrae probe concentration was assigned to red marrow, following the European Association of Nuclear Medicine (EANM) guidelines, as proposed in [29]. Finally, the value assigned for the remainder of the body was calculated as the difference from the total activity at a time-point minus the activity accounted in all organs, including the excreted one [25]. Total organ absorbed doses were then calculated using the ICRP 89 male and female phantoms from each subject. Data from each phantom across subjects were averaged together to create the average male and female phantom estimated doses.

### Statistical analysis

All statistical tests were run on the *R* software (v.4.2.3). All data was tested for normality using the Shapiro-Wilk test. Comparisons between female and male data and between the subgroups of 24h study vs. 2h study were performed using a Paired *t*-test, when data followed a normal distribution, or the Wilcoxon rank-sum test otherwise.

## Results

### Safety

No significant changes in vital signs, autonomic measures, or ECGs were observed after injection of [^64^Cu]FBP8. Subjects reported no discomfort after injection. Most lab values were in the normal range for all subjects. In one of the volunteers, hepatic transaminases were mildly increased, but the subject’s enzyme levels were chronically elevated for many years prior to probe injection and thus were not related to probe injection. In another volunteer, the PTT appeared to be elevated after probe injection, however, this was suspected to arise from residual heparin from the flush of the line. When the PTT was remeasured after the addition of heparinase, the PTT value returned to normal, confirming that the elevated PTT was due to heparin contamination and was a spurious result.

### Pharmacokinetics and Biodistribution

The mean radioactivity injected was 398 ± 136 MBq, (range 209 – 528). Figure 1 shows a typical biodistribution of [^64^Cu]FBP8 radiotracer over time.

**Fig. 1:**
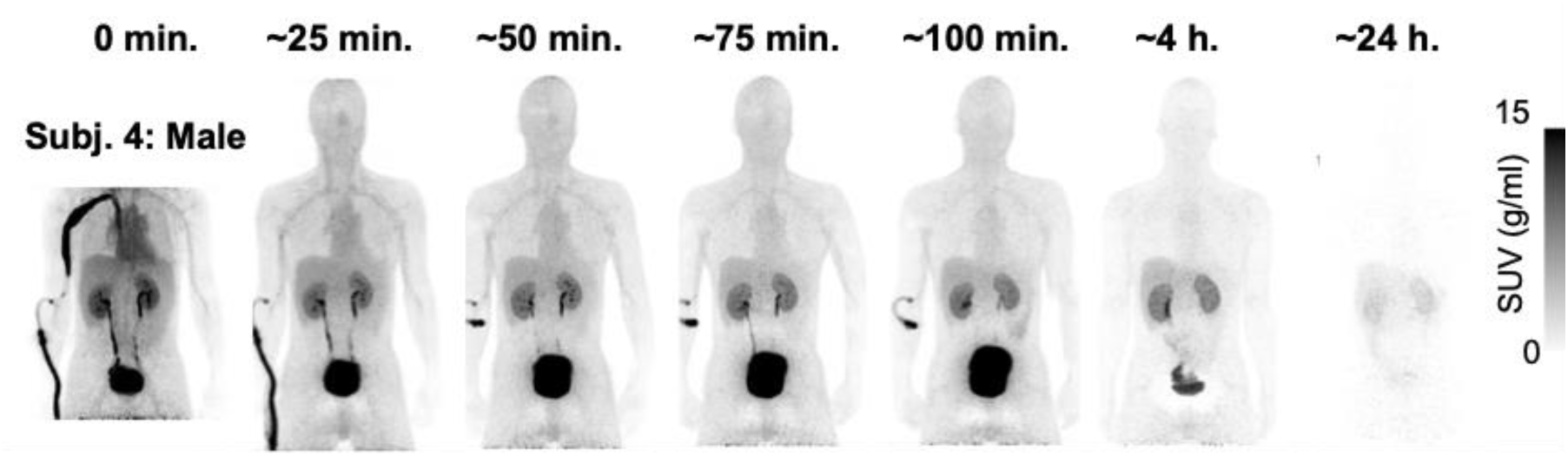
Representative biodistribution of [^64^Cu]FBP8 in the whole-body of a male subject from injection and up to 24 h. post injection.

Table 2 lists the SUV_mean_ values at 90 and 240 min post injection for various tissues for both females and males. No statistical differences were observed between female and male values: Wilcoxon Rank Sum Test results of *p*>0.05 for all ROIs.

**Table 2:** SUV_mean_ values at 90 and 240 min post injection for several organs across all subjects for both Females and Males. Data as Median [min – max].

| ROI | $\text{SUV}_{\text{mean}}^{90\text{min}}$ | | | $\text{SUV}_{\text{mean}}^{240\text{min}}$ | | |
| --- | --- | --- | --- | --- | --- | --- |
|  | All | Female | Male | All | Female | Male |
| Skeletal | 0.26 | 0.25 | 0.29 | 0.11 | 0.13 | 0.09 |
| Muscle | [0.08 0.40] | [0.23 0.26] | [0.08 0.40] | [0.06 0.16] | [0.11 0.16] | [0.06 0.11] |
| Left | 0.80 | 1.04 | 0.77 | 0.31 | 0.37 | 0.25 |
| Ventricle | [0.56 1.08] | [1.00 1.08] | [0.56 0.89] | [0.22 0.41] | [0.34 0.41] | [0.22 0.29] |
| Brain | 0.07 | 0.07 | 0.07 | 0.03 | 0.02 | 0.05 |
|  | [0.04 0.11] | [0.06 0.07] | [0.04 0.11] | [0.02 0.06] | [0.02 0.02] | [0.03 0.06] |
|  |  | 0.71 |  |  | 0.46 |  |
| Breast | N/A | [0.55 0.88] | N/A | N/A | [0.42 0.51] | N/A |
|  | 5.08 | 5.82 | 5.08 | 3.12 | 3.94 | 2.99 |
| Kidney | [3.25 7.44] | [4.21 7.44] | [3.25 5.83] | [2.66 4.94] | [2.94 4.94] | [2.66 3.31] |
| Large | 0.51 | 0.46 | 0.56 | 0.27 | 0.73 | 0.22 |
| Intestine | [0.38 0.66] | [0.45 0.47] | [0.38 0.66] | [0.08 1.25] | [0.20 1.25] | [0.08 0.35] |
|  | 0.79 | 0.83 | 0.63 | 0.29 | 0.34 | 0.26 |
| Myocardium | [0.54 1.04] | [0.79 0.87] | [0.54 1.04] | [0.23 0.38] | [0.30 0.38] | [0.23 0.29] |
|  | 1.49 | 1.51 | 1.35 | 1.02 | 1.09 | 0.95 |
| Liver | [1.20 2.71] | [1.49 1.53] | [1.20 2.71] | [0.91 1.13] | [1.05 1.13] | [0.91 0.99] |
|  | 0.25 | 0.25 | 0.20 | 0.09 | 0.10 | 0.08 |
| Lung | [0.16 0.30] | [0.25 0.25] | [0.16 0.30] | [0.07 0.11] | [0.09 0.11] | [0.07 0.10] |
|  | 0.80 | 0.77 | 0.81 | 0.47 | 0.47 | 0.52 |
| Pancreas | [0.55 0.92] | [0.73 0.80] | [0.55 0.92] | [0.38 0.66] | [0.41 0.52] | [0.38 0.66] |
|  | 0.73 | 0.67 | 0.73 | 0.54 | 0.83 | 0.52 |
| SI | [0.57 1.12] | [0.62 0.71] | [0.57 1.12] | [0.35 1.26] | [0.41 1.26] | [0.35 0.68] |
|  | 0.45 | 0.43 | 0.66 | 0.17 | 0.25 | 0.15 |
| Red Marrow | [0.14 0.75] | [0.40 0.45] | [0.14 0.75] | [0.14 0.31] | [0.19 0.31] | [0.14 0.16] |
|  | 0.52 | 0.52 | 0.52 | 0.24 | 0.24 | 0.24 |
| Spleen | [0.43 0.67] | [0.51 0.54] | [0.43 0.67] | [0.18 0.31] | [0.18 0.31] | [0.19 0.29] |
|  | 0.66 | 0.77 | 0.64 | 0.28 | 0.28 | 0.27 |
| Stomach | [0.33 0.95] | [0.75 0.79] | [0.33 0.95] | [0.19 0.35] | [0.21 0.34] | [0.19 0.35] |

The time-dependent biodistribution data (Figure 1 and Table 2) demonstrate rapid renal clearance with some uptake seen in the liver and the biliary tract, and very reduced background uptake in organs such as the lungs, brain or heart. Figure 2a shows the %ID/ml TACs for selected organs: lungs, liver, kidney, spleen and myocardium, further demonstrating the fast clearance of the probe and the low background activity in healthy regions. Figures 2b-c show the combined [^64^Cu]FBP8 radiotracer activity for all subjects, in %ID/ml, of the blood pool, showing a fast clearance. Fitting of individual subject data to a biexponential function shows a median distribution half-life of 4 min (range 2 – 11) and a median elimination half-life of 67 min (range 66 – 71). These results are consistent with a probe that has predominantly renal elimination with low protein binding and extracellular distribution.

**Fig. 2:**
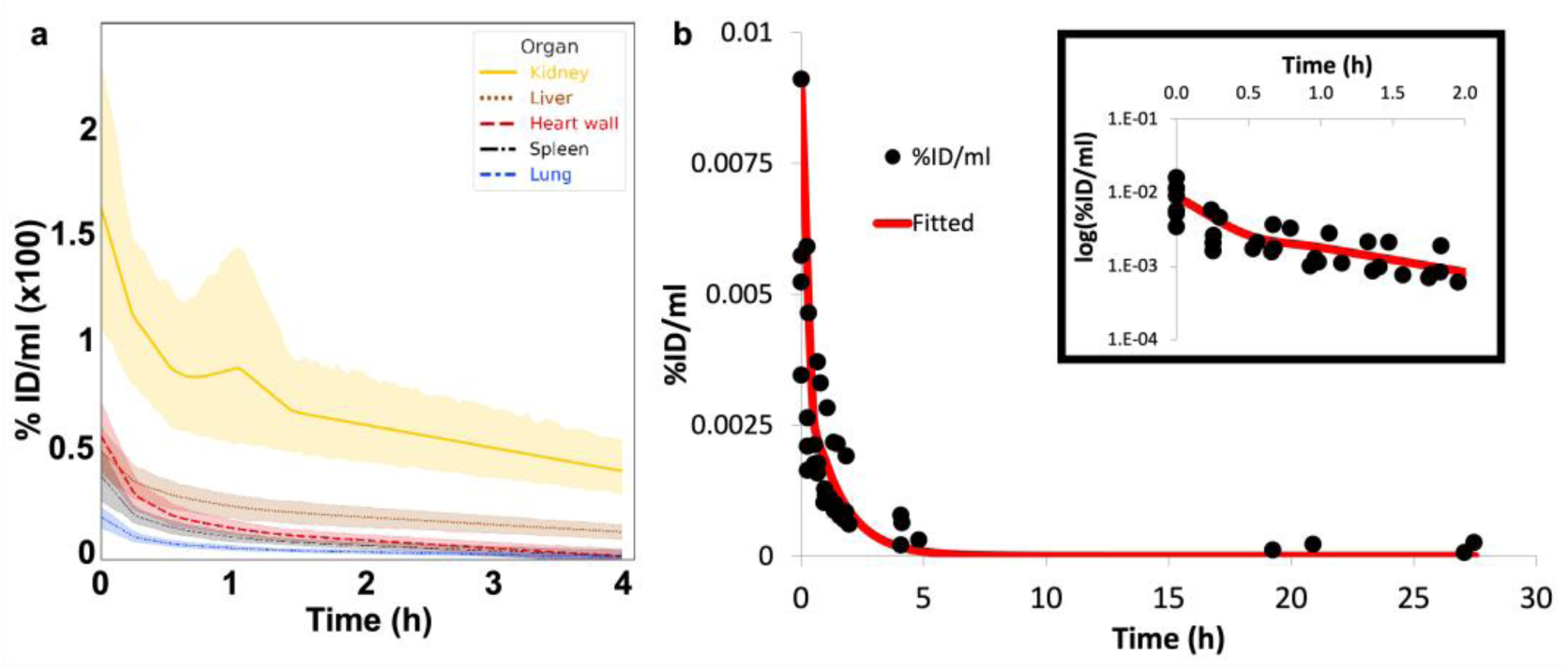
(**a**) Time activity curves (TAC) in %ID/ml (x100) showing uptake across subjects for some of the largest uptake ROIs; (**b**) Blood pool TAC across all subjects showing the fast blood clearance of [^64^Cu]FBP8. (**c**) Insert of (**b**) shows data 0-2h in a semi-log scale to highlight the bi-exponential nature of the curve.

Figure 3 shows an example of the data fitting across organs for one of the subjects (Subject 1, female) with mono- and bi-exponential functions. Fitting the total urine content obtained the following excretion fractions: 51% [43% - 66%]; and urine excretion half-lives: 26 min [18 min – 48 min]. These values were used for the Bladder model in OLINDA to estimate doses for the bladder [28].

**Fig. 3:**
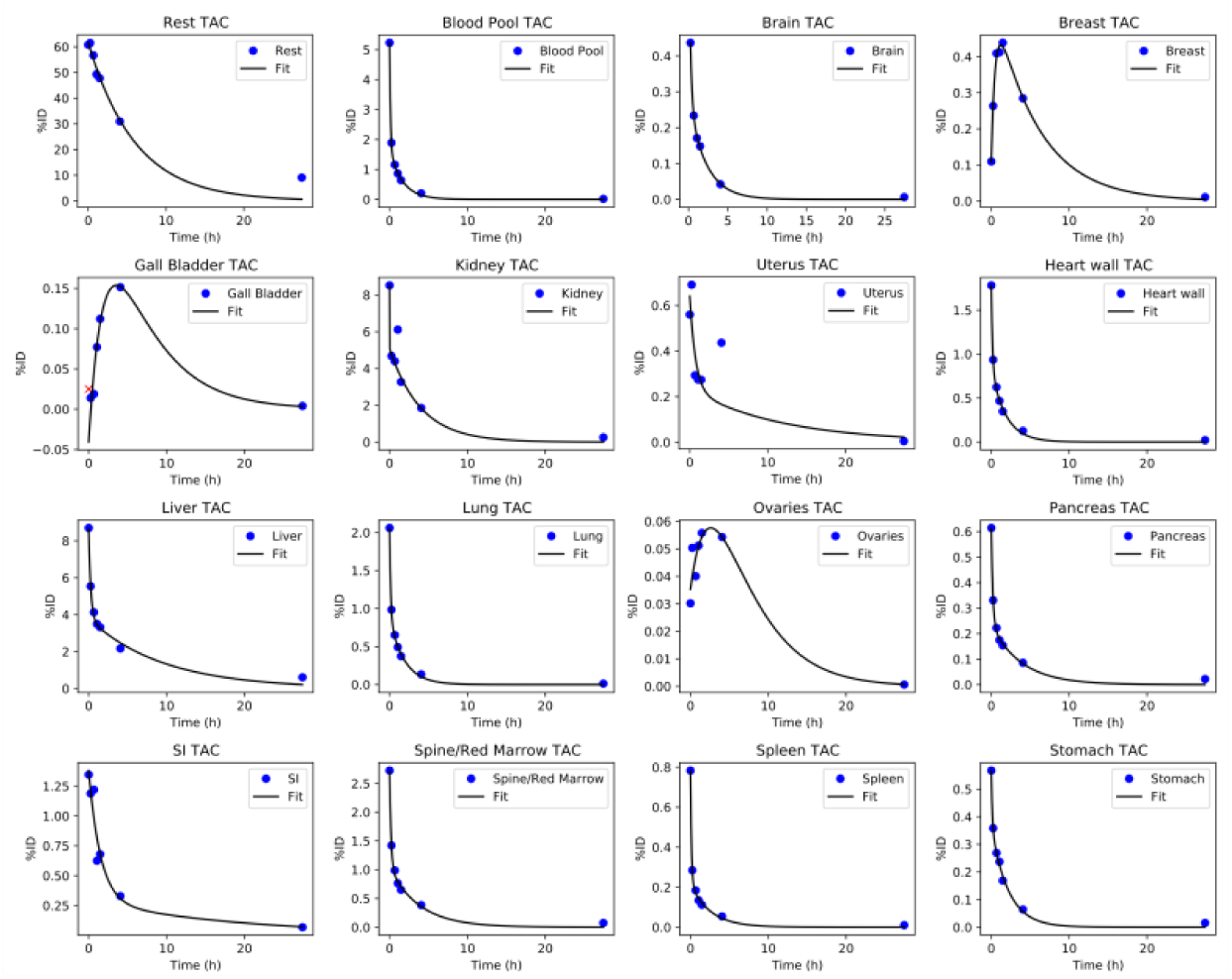
Examples of ROI fitting with single and double exponentials for a representative subject. *Rest*: Reminder of the Body; *SI* = Small Intestine.

TIAC values per organ are shown in Supplementary Table 1. We observed large differences in the TIAC values across all organs between those subjects for whom data was available out to 24 h post-injection vs. those for whom only data out to 2 h was available: TIACs from the 2 h only study were on average about 50% lower than those from the 24 h study (Supplementary Table 1), and accordingly calculated organ absorbed doses were also significantly lower for the 2 h only study compared to those from the 24 h study, except for sex-specific ROIs ovaries, prostate and testes (Supplementary Table 2 and Supplementary Figure 1). This analysis indicated the importance of collecting data at later time points. To avoid any potential dosimetry bias, final dosimetry results included in this study were obtained uniquely using those subjects undergoing the full 24h study.

Table 3 shows the organ absorbed doses (mGy/MBq) and the estimated effective doses (mSv/MBq) for a 1 hour voiding cycle on the bladder model in OLINDA, for the subjects having completed the full 24h study. The organs with the higher doses for both the male and female phantoms were: urinary bladder (0.075 and 0.091 mGy/MBq for male and female respectively), kidneys (0.050 and 0.056 mGy/MBq respectively), and liver (0.027 and 0.035 mGy/MBq respectively). The combined mean effective dose for males and females was 0.016 ± 0.0029 mSv/MBq. Females had higher organ doses and effective doses compared to males for all ROIs (Table 3). However, males and females did not show any significant differences on the TIACs (Supplementary Table 3).

**Table 3:** Organ absorbed doses (in mGy/MBq) and Effective Doses (in mSv/MBq) for the standard Male and Female phantoms (ICRP 89) using ICRP-103 standard for organ weights. Data shown as: mean (SD). N/A = not applicable. ^1^Paired *t*-test.

| Target Organ | Organ Doses (mGy/MBq) |  | *p*-value<sup>1</sup> |
| --- | --- | --- | --- |
|  | Male phantom | Female phantom |  |
| Adrenals | **0.017 (0.0016)** | **0.020 (0.0019)** | **<0.01** |
| Brain | **0.0025 (0.0001)** | **0.0032 (0.0002)** | **<0.001** |
| Breasts | **N/A** | **0.011 (0.0035)** | **N/A** |
| Esophagus | **0.0134 (0.0019)** | **0.016 (0.0021)** | **<0.001** |
| Eyes | **0.0121 (0.0022)** | **0.0148 (0.0027)** | **<0.01** |
| Gallbladder Wall | **0.022 (0.0048)** | **0.026 (0.0060)** | **<0.01** |
| Left colon | **0.018 (0.0028)** | **0.021 (0.0034)** | **<0.01** |
| Small Intestine | **0.022 (0.0030)** | **0.026 (0.0037)** | **<0.01** |
| Stomach Wall | **0.015 (0.0020)** | **0.018 (0.0027)** | **<0.01** |
| Right colon | **0.016 (0.0022)** | **0.019 (0.0029)** | **<0.01** |
| Rectum | **0.016 (0.0026)** | **0.021 (0.0031)** | **<0.001** |
| Heart Wall | **0.010 (0.0016)** | **0.013 (0.0018)** | **<0.001** |
| Kidneys | **0.050 (0.011)** | **0.056 (0.012)** | **<0.01** |
| Liver | **0.027 (0.0172)** | **0.035 (0.0218)** | **0.046** |
| Lungs | **0.0055 (0.0004)** | **0.0072 (0.0005)** | **<0.001** |
| Testes / Ovaries | **0.011 (0.0050)** | **0.025 (0.0154)** | **N/A** |
| Pancreas | **0.015 (0.0073)** | **0.019 (0.0088)** | **0.016** |
| Prostate / Uterus | **0.017 (0.003)** | **0.030 (0.012)** | **N/A** |
| Salivary Glands | **0.0133 (0.0024)** | **0.016 (0.0029)** | **<0.01** |
| Red Marrow | **0.013 (0.0016)** | **0.016 (0.0020)** | **<0.001** |
| Osteogenic Cells | **0.0140 (0.0021)** | **0.0145 (0.0022)** | **<0.01** |
| Spleen | **0.0081 (0.00086)** | **0.010 (0.0010)** | **<0.001** |
| Thymus | **0.0132 (0.0021)** | **0.016 (0.0027)** | **<0.01** |
| Thyroid | **0.0135 (0.0024)** | **0.016 (0.0028)** | **<0.01** |
| Urinary Bladder Wall | **0.075 (0.0040)** | **0.091 (0.0052)** | **<0.001** |
| Total Body | **0.015 (0.0020)** | **0.018 (0.0023)** | **<0.001** |
| **Effective Dose (mSv/MBq)** | **0.013 (0.0009)** | **0.018 (0.0015)** | **<0.001** |

To further explore the impact radioisotope choice on dosimetry, we simulated the effect of replacing Cu-64 in [^64^Cu]FBP8 with Ga-68 by first decay-correcting the original Cu-64 data, and then decay-uncorrecting it using the Ga-68 half-life. From this simulated data, the estimated mean effective dose for a putative [^68^Ga]FBP8 probe would be higher than that of [^64^Cu]FBP8 (0.020±0.0031 vs. 0.0016±0.0029, *p*<0.001, Supplementary Table 4).

## Discussion

This report describes the safety, whole body pharmacokinetics, and dosimetry of [^64^Cu]FBP8, and has several important findings. First, in a group of 8 healthy subjects, there were no adverse events related to the probe [^64^Cu]FBP8.

The whole body distribution and pharmacokinetics of [^64^Cu]FBP8 were similar to that predicted from rodent models. [^64^Cu]FBP8 showed an extracellular distribution and was cleared from circulation rapidly, with a distribution half-life of 4 min and an elimination half-life of about 70 min consistent with glomerular filtration. Interestingly, in those subjects in whom the peripheral IV line was flushed with normal saline, small thrombi formed quite rapidly in the IV catheter. This was not seen when the line was flushed with heparinized saline, underscoring the need for an anticoagulant-containing flush to be used with indwelling IV catheters. Apart from the excretory organs, the background signal with [^64^Cu]FBP8 was quite low. This distribution suggests that [^64^Cu]FBP8 PET may be sensitive for detecting fibrin in the thorax, the neck, the brain, and peripheral vessels. Since blood and background signal decreases with time, delayed imaging may be preferable.

Because not all subjects completed the three scan sessions, we tested the effect of estimating TIACs (and doses) from the data fitted using the first two hours of data collection post injection vs. using the 24 hours. We observed lower TIACs (and doses) on subjects with 2-hour data collection vs. those with 24h collection (Supplementary Table 1 and 2). These results suggests that a shorter data collection period (∼2h) with Cu-64 dosimetry studies is not sufficient as it leads to underestimation of the dosimetry values. Our data supports the idea that data collection closer to 2 half-lives (∼24h for Cu-64) is a reasonable timing to avoid a bias towards lower dosimetry estimates due to poor curve fitting to obtain TIACs. This is the first study that presents data to support the importance of imaging timing in dosimetry bias. Therefore, to avoid such bias, final dosimetry results on this study used only the subjects who completed the 24h imaging study.

The organs with the highest doses were consistent with a renal clearance probe: the urinary bladder received the highest dose, followed by the kidneys and then the liver. Absorbed doses were higher for females vs. males, a common finding in most dosimetry studies [22, 27, 30–31]. However, neither the SUV values at 90 or 240 min (Table 2), nor the TIACs (Supplementary Table 3) were significantly different between sexes. These results suggest that the higher *S* values per organ for the female phantom, a result of the smaller female organ and body size [32], is responsible for the sex-related difference in doses observed, as we have previously noted [33].

The combined mean effective dose across both male and female phantom is 0.016 mSv/MBq with a 1h voiding model and 0.020 mSv/MBq with a 2h voiding model, which is similar to that of the widely used [^18^F]fluorodeoxyglucose ([^18^F]FDG, 0.020mSv/MBq) [34]. Our results calculated from human data are in accord with predicted dosimetry values extrapolated from a rat model: 0.021 mSv/MBq and 0.027 mSv/MBq for the male and female phantom respectively using a 1.5 h bladder model in a prior version of OLINDA (v.1.0) [18]. This study confirms the predicted dosimetry estimates and confirms the low effective doses for human use compared to other probes (Table 4). [^64^Cu]FBP8 has more favorable dosimetry values compared to other ^64^Cu-based PET probes and similar values compared to ^68^Ga-based PET probes (Table 4 and [33]), despite the 11-fold longer half-life of Cu-64 compared to Ga-68. This favorable dosimetry is a consequence of predominantly extracellular distribution and fast renal elimination. Additionally, the higher positron energy of Ga-68 outweighs the longer half-life of Cu-64 in terms of absorbed dose (Supplementary Table 4). Additionally, the longer half-life of Cu-64 can provide advantages in certain clinical situations, such as imaging the next day to evaluate whether a thrombus has resolved. The longer half-life is also advantageous for shipping the probe over long distances to other clinical sites.

**Table 4:** Comparison of [^64^Cu]FBP8 with Mean Effective Doses of other Cupper-64 based PET Probes and [^18^F]FDG. In square brackets: the range of mean effective doses reported.

| Compound | Effective Doses<br>(mSv/MBq) | Voiding<br>model | Reference |
| --- | --- | --- | --- |
| [<sup>64</sup>Cu]CuCl<sub>2</sub> | 0.056 [0.051 – 0.061] in<br>healthy controls | Unspecified | [30] |
| [<sup>64</sup>Cu]NODAGA-RGDyK | [0.029 – 0.038] | 5h | [35] |
| [<sup>64</sup>Cu]DOTATATE | 0.032 | 2h | [25, 31] |
| [<sup>64</sup>Cu]ATSM | 0.036 | No voiding | [26] |
| [<sup>64</sup>Cu]LLP2A | 0.036 ± 0.0006<br>[0.029 – 0.046] | 2h | [27] |
| [<sup>64</sup>Cu]DOTA-AE105 | 0.0284 [0.025 – 0.032] | Unspecified | [31] |
| [<sup>68</sup>Ga]NODAGA-MJ9 | [0.018 – 0.023] | 30 min & 1 h | [36] |
| [<sup>68</sup>Ga]DOTA-E-[c(RGDfK)]<sub>2</sub> | [0.017 – 0.024] | 1 h | [32] |
| [<sup>18</sup>F]FDG | 0.020 [0.013 – 0.029] | Unspecified | [34] |
| [<sup>68</sup>Ga]CBP8 | 0.018 [0.016 – 0.021] | 1 h | [33] |
| **[<sup>64</sup>Cu]FBP8** | **0.016 [0.013 – 0.020]** | **1 h** | **This study** |
| **[<sup>64</sup>Cu]FBP8** | **0.019 [0.016 – 0.024]** | **2 h** | **This study** |

This dosimetry study suggests that in order to avoid reaching the maximum allowed dose on any organ established by the Radioactive Drug Research Committee (RDRC) guidelines (5 cGy), the maximum radiotracer concentrations that could be clinically injected are ∼700 MBq for males and ∼550 MBq for females.

Our study has several limitations. First, despite recruiting 8 subjects for this study, only 4 were included in the final dosimetry results: 1 subject had a scanner failure during acquisition, and 3 subjects declined to continue with the additional imaging sessions at 4h and 24h. Nevertheless, the data obtained from the remaining subjects have provided strong evidence of the dosimetry estimates regarding organ and effective doses, and it has allowed us to investigate the impact of scanning duration in dosimetry estimates for the first time. Second, our study only included healthy subjects. Further studies with patients, in particular those with chronic kidney disease where the rapid renal clearance of the probe may be slowed could affect the dosimetry estimates.

In summary, the favorable dosimetry estimates, comparable to other commonly used probes in the nuclear medicine field, make [^64^Cu]FBP8 an attractive clinical candidate for non-invasive fibrin detection and quantification. The low background signal in vascular territories important for thrombus detection suggests a high potential for accurate diagnoses using [^64^Cu]FBP8 PET. Low PET probe doses are of extreme importance, above all in longitudinal studies, which is often the case of cardiovascular patients [22], or in oncological or neurological cases. The rapid clearance of the probe, together with the long half-life of the tracer, is ideal for clinical situations, where a probe can be injected and patients can be imaged either within a short time-window after injection, or several hours later, even the next day.

## Conclusion

This study has demonstrated fast blood clearance and renal excretion of [^64^Cu]FBP8 as well as similar or lower dosimetry values compared to other clinically used probes. These data demonstrate the potential of [^64^Cu]FBP8 as a very favorable PET probe for a myriad of potential clinical applications where fibrin is an important driver/feature, such as most cardiovascular applications (stroke, atherosclerosis, myocardial infarction, atrial fibrillation, etc.) as well as other non-cardiovascular ones, such as tumors, lung fibrosis or even neuro-degenerative studies, areas where our group is actively working.

## Author contributions

DIG and PC contributed to the study concepts and design. DIG, ALP and DES contributed to the data acquisition. PD and PC contributed to the probe conception and design. DIG and CC contributed to PET image reconstruction. DIG contributed to the data analysis and statistical analysis. DIG and PC contributed to the manuscript preparation. All authors contributed to manuscript review and read and approved the final manuscript.

## Ethical Approval

All procedures performed in studies involving human participants were in accordance with the ethical standards of the institutional and/or national research committee and with the 1964 Helsinki declaration and its later amendments or comparable ethical standards.

## Data Availability

All research data are available from the corresponding author upon request.

