## Supplemental Material for "Dosimetry of [^64^Cu]FBP8: a fibrin-binding PET probe"

### Electronic Supplementary Material

**Journal: Molecular Imaging and Biology**

Authors: David Izquierdo-Garcia[1,2,3,4], Pauline Désogère[1,5], Anne L. Philip[6], David E. Sosnovik[1,5,6,7], Ciprian Catana[1,2,5], Peter Caravan[1,2,5]

[1]Athinoula A. Martinos Center for Biomedical Imaging, Department of Radiology, Massachusetts General Hospital, Boston, MA

[2]Harvard Medical School, Boston, MA

[3]Harvard-MIT Division of Health Sciences and Technology, Cambridge, MA

[4]Bioengineering Department, Universidad Carlos III, Madrid, Spain

[5]Institute for Innovation in Imaging, Massachusetts General Hospital, Boston, MA

[6]Cardiovascular Research Center, Cardiology Division, Dept. of Medicine, Massachusetts General Hospital and Harvard Medical School, Boston MA

[7]Cardiology Division, Department of Medicine, Massachusetts General Hospital and Harvard Medical School, Boston, MA.

**Corresponding author**:

David Izquierdo-Garcia

149 Thirteen St. Suite 2301,

Charlestown, MA, 02129, USA

**Supplementary Material and Methods**

**Dosimetry: OLINDA Bladder model**

Since the probe is mainly excreted into the urine, the bladder wall is one of the main dose receptors. The total volume of the bladder and its total organ uptake was calculated using an in-house script that segmented the whole bladder (Matlab, Mathworks, Natick, MA). TIACs for the urinary bladder were calculated using the voiding model on the OLINDA software in a similar way as explained in [27]. Briefly, this model allows the use of up to 4 different excretion fractions of the urinary bladder (with their corresponding biological half-lives) as well as choosing a voiding interval, i.e. the time for a typical patient to void the bladder in a clinical setting. Since subjects were asked to void their bladder during the break between the 2h and 4h scans, the voiding volume was estimated using the whole content of the bladder on the last imaging time-point prior to voiding. Similarly, the voiding volume between the 4h and 24h post injection imaging sessions was estimated from the bladder volume at 4h post injection. The total urine content was then calculated by adding at each time point the voided volume and the bladder volume. This curved was fitted into an inverse exponential (see equation below) using the GRG Solver in Excel 2013 as explained in [27]:

$$Total\ Urine\ Content = A \cdot \left(1 - e^{-\lambda t}\right),$$

where $A$ is the excretion fraction of the compartment model (exponential) and $\lambda$ is the rate constant that defines the urine excretion half-life as Ln(2)/$\lambda$. $A$ values were restricted to be between 0 and 100, defining the percentage of the total injected dose excreted through urine.

**Supplementary Results**

**Supplementary Table 1**: Comparison of the organ time-integrated activity coefficients (TIAC, MBq-h/MBq) for the subjects undergoing the full 24h study vs. the limited 2h study. Sex-specific ROIs (Breast, Ovaries, Testes, Uterus and Prostate) have been removed from the list since only males were included on the 2h study. Data presented as Median [Min – Max]. $RC(\%) = Relative\ change\ (\%) = 100 \times (\overline{TIAC_{24h}} - \overline{TIAC_{2h}})/\overline{TIAC_{24h}}$.
[1]Paired *t*-test

| | TIAC (MBq-h/MBq) | | | |
|---|---|---|---|---|
| | **24h Study (N=4)** | **2h study (N=3)** | ***RC (%)*** | ***p-value*[1]** |
| **Blood Pool** | 0.030 [0.018 - 0.034] | 0.013 [0.009 - 0.036] | 31.20 | 0.34 |
| **Brain** | 0.0092 [0.0073 - 0.017] | 0.0075 [0.0055 - 0.008] | 33.65 | 0.24 |
| **Gall Bladder** | 0.0063 [0.0043 - 0.0153] | 0.0004 [0.0003 - 0.0007] | 93.75 | **<0.05** |
| **Kidney** | 0.19 [0.113 - 0.20] | 0.10 [0.082 - 0.13] | 41.34 | **<0.05** |
| **Large Intestine** | 0.0048 [0.0032 - 0.0060] | 0.0044 [0.0042 - 0.0046] | 5.76 | 0.71 |
| **Heart Wall** | 0.016 [0.011 - 0.017] | 0.012 [0.012 - 0.013] | 17.82 | 0.20 |
| **Liver** | 0.33 [0.254 - 1.00] | 0.21 [0.091 - 0.31] | 57.19 | 0.26 |
| **Lung** | 0.018 [0.014 - 0.021] | 0.013 [0.010 - 0.013] | 35.10 | **<0.05** |
| **Pancreas** | 0.0125 [0.0076 - 0.0357] | 0.0058 [0.0035 - 0.0060] | 70.06 | 0.18 |
| **Small Intestine** | 0.069 [0.029 - 0.080] | 0.023 [0.017 - 0.023] | 66.31 | **<0.05** |
| **Red Marrow** | 0.046 [0.023 - 0.074] | 0.034 [0.010 - 0.052] | 31.49 | 0.40 |
| **Spleen** | 0.0052 [0.0049 - 0.0063] | 0.0030 [0.0029 - 0.0046] | 34.91 | **<0.05** |

| | | | | |
|---|---|---|---|---|
| **Stomach** | 0.0071<br>[0.0059 - 0.0082] | 0.0035<br>[0.0022 - 0.0067] | 41.43 | 0.066 |
| **Reminder of Body** | 8.90<br>[6.57 - 10.20] | 2.34<br>[1.84 - 2.63] | 73.72 | **<0.05** |

**Supplementary Table 2:** Average organ absorbed doses across the male and female phantoms (ICRP 89, in mGy/MBq) and Effective Doses (in mSv/MBq) comparing data from subjects that completed the full 24h study vs. those who only completed the initial 2h. Data shown as: mean (SD). N/A = not applicable. [1]Wilcoxon rank-sum test. [2]Unpaired t-test.

| | **Organ Doses (mGy/MBq)** | | |
|---|---|---|---|
| **Target Organ** | Average (SD) 24h Study (n=4) | Average (SD) 2h study (n=3) | *p-value* |
| Adrenals | **0.019 (0.0022)** | **0.006 (0.0011)** | **$^{1}p<0.001$** |
| Brain | **0.003 (0.0004)** | **0.001 (0.0001)** | **$^{1}p<0.001$** |
| Breasts | **0.011 (0.0035)** | **0.004 (0.0007)** | **$^{2}p<0.05$** |
| Esophagus | **0.015 (0.0023)** | **0.004 (0.0009)** | **$^{1}p<0.001$** |
| Eyes | **0.013 (0.0027)** | **0.004 (0.0007)** | **$^{1}p<0.001$** |
| Gallbladder Wall | **0.024 (0.0054)** | **0.006 (0.0010)** | **$^{2}p<0.001$** |
| Left colon | **0.019 (0.0033)** | **0.007 (0.0009)** | **$^{2}p<0.001$** |
| Small Intestine | **0.024 (0.0038)** | **0.007 (0.0008)** | **$^{1}p<0.001$** |
| Stomach Wall | **0.017 (0.0028)** | **0.005 (0.0009)** | **$^{2}p<0.001$** |
| Right colon | **0.017 (0.0029)** | **0.005 (0.0009)** | **$^{1}p<0.001$** |
| Rectum | **0.019 (0.0038)** | **0.006 (0.0015)** | **$^{2}p<0.001$** |
| Heart Wall | **0.012 (0.0023)** | **0.007 (0.0024)** | **$^{2}p<0.01$** |
| Kidneys | **0.053 (0.0112)** | **0.030 (0.0063)** | **$^{2}p<0.001$** |
| Liver | **0.031 (0.0186)** | **0.013 (0.0060)** | **$^{1}p<0.01$** |
| Lungs | **0.006 (0.0010)** | **0.002 (0.0005)** | **$^{2}p<0.001$** |
| Ovaries | **0.025 (0.0154)** | **0.006 (0.0007)** | $^{1}p=0.06$ |
| Pancreas | **0.017 (0.0077)** | **0.005 (0.0009)** | **$^{1}p<0.001$** |
| Prostate | **0.017 (0.0028)** | **0.014 (0.0056)** | $^{2}p=0.37$ |
| Salivary Glands | **0.014 (0.0027)** | **0.004 (0.0007)** | **$^{1}p<0.001$** |
| Red Marrow | **0.015 (0.0024)** | **0.005 (0.0006)** | **$^{1}p<0.01$** |
| Osteogenic Cells | **0.014 (0.0020)** | **0.004 (0.0002)** | **$^{1}p<0.001$** |

| | | | |
|---|---|---|---|
| Spleen | **0.009 (0.0013)** | **0.004 (0.0007)** | **[2]*p*<0.001** |
| Testes | **0.011 (0.0050)** | **0.007 (0.0041)** | [2]*p*=0.36 |
| Thymus | **0.015 (0.0028)** | **0.004 (0.0008)** | **[2]*p*<0.001** |
| Thyroid | **0.015 (0.0027)** | **0.004 (0.0007)** | **[1]*p*<0.01** |
| Urinary Bladder Wall | **0.083 (0.0098)** | **0.071 (0.0105)** | **[2]*p*<0.05** |
| Uterus | **0.030 (0.0119)** | **0.008 (0.0006)** | **[2]*p*<0.05** |
| Total Body | **0.016 (0.0027)** | **0.005 (0.0009)** | **[1]*p*<0.001** |
| **Effective Dose (mSv/MBq)** | **0.016 (0.0029)** | **0.007 (0.0010)** | **[2]*p*<0.001** |

**Supplementary Table 3**: Values for the organ time-integrated activity coefficients (TIAC, MBq-h/MBq). Data presented as Median [Min – Max]. [1]Wilcoxon Rank Sum Test

| | **TIAC (MBq-h/MBq)** | | |
|---|---|---|---|
| | **Female** | **Male** | ***p-value*[1]** |
| **Blood Pool** | 0.030 [0.028 - 0.032] | 0.026 [0.0176 - 0.034] | 1 |
| **Brain** | 0.0086 [0.0073 - 0.0099] | 0.0126 [0.0085 - 0.0167] | 0.67 |
| **Breast** | 0.035 [0.031 - 0.039] | N/A | N/A |
| **Gall Bladder** | 0.010 [0.0043 - 0.015] | 0.0063 [0.0062 - 0.0063] | 1 |
| **Kidney** | 0.20 [0.19 - 0.20] | 0.15 [0.11 - 0.19] | 0.33 |
| **Large Intestine** | 0.0048 [0.0047 - 0.0049] | 0.0046 [0.0032 - 0.0060] | 1 |
| **Heart Wall** | 0.016 [0.015 - 0.017] | 0.014 [0.011 - 0.017] | 1 |
| **Liver** | 0.31 [0.25 - 0.37] | 0.64 [0.28 - 1.00] | 0.67 |
| **Lung** | 0.018 [0.018 - 0.018] | 0.018 [0.014 - 0.021] | 1 |
| **Ovaries / Prostate** | 0.0034 [0.0011 - 0.0057] | 0.0020 [0.0012 - 0.0028] | N/A |
| **Pancreas** | 0.0087 [0.0076 - 0.0099] | 0.0254 [0.0151 - 0.0357] | 0.33 |
| **Small Intestine** | 0.076 [0.073 - 0.080] | 0.047 [0.029 - 0.065] | 0.33 |
| **Red Marrow** | 0.059 [0.043 - 0.074] | 0.035 [0.023 - 0.048] | 0.67 |
| **Spleen** | 0.0056 [0.0049 - 0.0063] | 0.0052 [0.0051 - 0.0054] | 1 |
| **Stomach** | 0.0079 [0.0076 - 0.0082] | 0.0063 [0.0059 - 0.0067] | 0.33 |
| **Uterus / Testes** | 0.031 [0.030 - 0.032] | 0.0010 [0.0006 - 0.0013] | N/A |
| **Reminder of Body** | 9.18 [8.17 - 10.20] | 8.10 [6.57 - 9.63] | 0.67 |

**Supplementary Table 4:** Average organ absorbed doses across the male and female phantoms (ICRP 89, in mGy/MBq) and Effective Doses (in mSv/MBq) comparing data from our study using the $^{64}$Cu radioisotope vs. the simulated $^{68}$Ga one. Data shown as: mean (SD). N/A = not applicable. [2]Wilcoxon rank-sum test. [1]Paired t-test.

| | **Organ Doses (mGy/MBq)** | | |
|---|---|---|---|
| **Target Organ** | Average (SD) 24h Study (n=4) | Average (SD) 2h study (n=3) | *p-value* |
| Adrenals | **0.019 (0.0022)** | **0.015 (0.0024)** | **[1]*p*<0.001** |
| Brain | **0.003 (0.0004)** | **0.002 (0.0004)** | **[1]*p*<0.001** |
| Breasts | **0.011 (0.0035)** | **0.009 (0.0020)** | [1]*p*=0.33 |
| Esophagus | **0.015 (0.0023)** | **0.009 (0.0011)** | **[1]*p*<0.001** |
| Eyes | **0.013 (0.0027)** | **0.008 (0.0010)** | **[1]*p*<0.001** |
| Gallbladder Wall | **0.024 (0.0054)** | **0.017 (0.0029)** | **[1]*p*<0.01** |
| Left colon | **0.019 (0.0033)** | **0.019 (0.0036)** | [1]*p*=0.65 |
| Small Intestine | **0.024 (0.0038)** | **0.022 (0.0039)** | [1]*p*=0.09 |
| Stomach Wall | **0.017 (0.0028)** | **0.013 (0.0015)** | **[1]*p*<0.001** |
| Right colon | **0.017 (0.0029)** | **0.010 (0.0012)** | **[1]*p*<0.001** |
| Rectum | **0.019 (0.0038)** | **0.016 (0.0041)** | **[1]*p*<0.05** |
| Heart Wall | **0.012 (0.0023)** | **0.023 (0.0047)** | **[1]*p*<0.001** |
| Kidneys | **0.053 (0.0112)** | **0.070 (0.0340)** | **[2]*p*<0.05** |
| Liver | **0.031 (0.0186)** | **0.022 (0.0034)** | **[2]*p*=0.64** |
| Lungs | **0.006 (0.0010)** | **0.007 (0.0010)** | **[1]*p*=0.17** |
| Ovaries | **0.025 (0.0154)** | **0.025 (0.0129)** | [1]*p*=0.99 |
| Pancreas | **0.017 (0.0077)** | **0.016 (0.0023)** | [2]*p*=0.74 |
| Prostate | **0.017 (0.0028)** | **0.016 (0.0039)** | [1]*p*=0.72 |
| Salivary Glands | **0.014 (0.0027)** | **0.008 (0.0009)** | **[1]*p*<0.001** |
| Red Marrow | **0.015 (0.0024)** | **0.010 (0.0012)** | **[1]*p*<0.001** |
| Osteogenic Cells | **0.014 (0.0020)** | **0.008 (0.0003)** | **[2]*p*<0.01** |
| Spleen | **0.009 (0.0013)** | **0.013 (0.0019)** | **[1]*p*<0.001** |

| | | | |
|---|---|---|---|
| Testes | **0.011 (0.0050)** | **0.009 (0.0016)** | [1]*p*=0.52 |
| Thymus | **0.015 (0.0028)** | **0.009 (0.0012)** | **[1]*p*<0.001** |
| Thyroid | **0.015 (0.0027)** | **0.008 (0.0009)** | **[1]*p*<0.001** |
| Urinary Bladder Wall | **0.083 (0.0098)** | **0.248 (0.0351)** | **[2]*p*<0.01** |
| Uterus | **0.030 (0.0119)** | **0.040 (0.0227)** | [1]*p*=0.21 |
| Total Body | **0.016 (0.0027)** | **0.011 (0.0015)** | **[1]*p*<0.001** |
| **Effective Dose (mSv/MBq)** | **0.016 (0.0029)** | **0.020 (0.0031)** | **[1]*p*<0.001** |

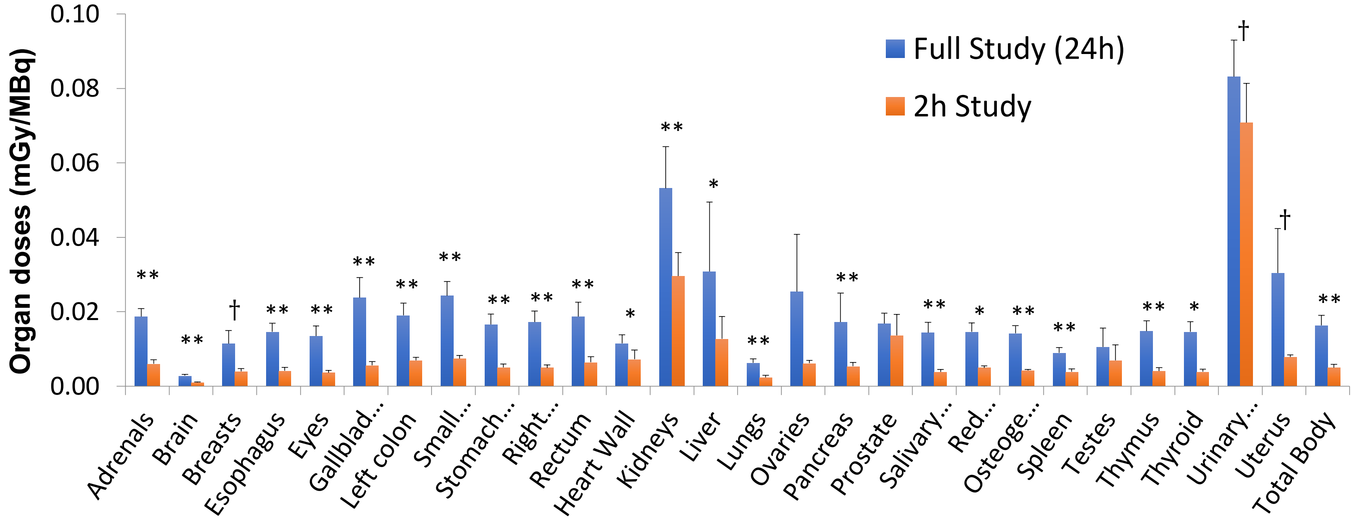


**Supplementary Figure 1**: Comparison of Organ doses (mGy/MBq) for the full study (24h) vs. the 2h study. All organs are significantly lower for the 2h study except for sex-specific ROIs Ovaries, Prostate and Testes. †: p<0.05; *: p<0.01; **: p<0.001.
